# *“Even things they won’t share with their sisters-in-law”* – Assessing an integrated Community Health Worker intervention on person-centered postpartum contraception in rural Nepal

**DOI:** 10.1101/2024.05.23.24307832

**Authors:** Nandini Choudhury, Wan-Ju Wu, Rekha Khatri, Aparna Tiwari, Aradhana Thapa, Samrachna Adhikari, Indira Basnett, Ved Bhandari, Aasha Bhatta, Bhawana Bogati, Laxman Datt Bhatt, David Citrin, Scott Halliday, Sonu Khadka, Yashoda Kumari Bhat Ksetri, Lal Bahadur Kunwar, Kshitiz Rana Magar, Nutan Marasini, Duncan Maru, Isha Nirola, Rashmi Paudel, Bala Rai, Ryan Schwarz, Sita Saud, Dikshya Sharma, Goma Niroula Shrestha, Ramesh Shrestha, Poshan Thapa, Hari Jung Rayamazi, Sheela Maru, Sabitri Sapkota

**Affiliations:** Department of Population Health, New York University Grossman School of Medicine, New York, NY, USA; Possible, New York, NY, USA; Arnhold Institute for Global Health, Icahn School of Medicine at Mount Sinai, New York, NY, USA; Department of Obstetrics, Gynecology, and Reproductive Science, Icahn School of Medicine at Mount Sinai, New York, NY, USA; Elmhurst Hospital, New York City Health + Hospitals, New York, NY, USA; Possible, Kathmandu, Nepal; Global Health and Development Program, Laney Graduate School, Emory University, Atlanta, GA, USA; Patan Academy of Medicine, Kathmandu, Nepal; Nyaya Health Nepal, Kathmandu, Nepal; Division of Community Health and Humanities, Memorial University of Newfoundland, St. John’s, Newfoundland, Canada; Department of Global Health, University of Washington, Seattle, WA, USA; Department of Anthropology, University of Washington, Seattle, WA, USA; Washington State Department of Children, Youth & Families, Olympia, WA, USA; Medic, Kathmandu, Nepal; Transcultural Psychosocial Organization Nepal, Kathmandu, Nepal; FrontBooth Solutions, Kathmandu, Nepal; Department of Internal Medicine, Icahn School of Medicine at Mount Sinai, New York, NY, USA; Boston University School of Public Health, Boston, MA, USA; Nursing and Social Security Division, Kathmandu, Nepal; Department of Medicine, Division of Global Health Equity, Brigham and Women’s Hospital, Boston, MA, USA; Department of Medicine, Harvard Medical School, Boston, MA, USA; Department of Medicine, Division of General Internal Medicine, Massachusetts General Hospital, Boston, MA, USA; Civil Service Hospital, Kathmandu, Nepal; Department of Health Service, Ministry of Health and Population, Kathmandu, Nepal; School of Health and Allied Sciences, Pokhara University, Pokhara, Nepal; School of Population and Global Health, McGill University, Quebec, Canada

## Abstract

Postpartum contraceptive counseling and access are challenging in Nepal’s remote, hilly areas, driving a disproportionately higher unmet need for contraception. Community health workers (CHWs) play an important role in delivering healthcare in difficult to reach places in Nepal, but there is limited evidence on the ideal CHW model and its impact over time. We implemented a pilot program in two rural districts in Nepal where full-time, salaried, and supervised CHWs delivered a bundled reproductive, maternal, newborn, and child health (RMNCH) intervention. This included a person-centered contraceptive counseling component adapted from the Balanced Counseling Strategy. Applying a type 2 hybrid effectiveness-implementation study approach, we conducted a non-randomized pre-post study with repeated measurements and nested qualitative data collection to assess the intervention’s reach, effectiveness, adoption, implementation, and maintenance. This paper describes the postpartum contraceptive outcomes associated with the integrated RMNCH intervention over a five-year period. Compared to the pre-intervention period, we observed a higher ward-level post-intervention postpartum contraceptive prevalence stratified by early postpartum (RR: 2.20; 95% CI: 1.96, 2.48) and late postpartum (RR: 1.70; 95% CI: 1.50, 1.93) periods, after adjusting for district and intervention site. Although we observed high rates of lactational amenorrhea method (LAM) in most intervention sites, the proportion of women who switched from LAM to another effective method was relatively low. Qualitative data indicate that CHWs’ longitudinal engagement enabled them to build trust with participants in their community, which likely contributed to their uptake of modern contraceptive methods. Barriers to modern contraceptive use included fear of side effects, limited autonomy for women, peer influence, and contraceptive unavailability. Implementation barriers included distance, challenging physical terrain, and increased travel times during the rainy season. This study contributes to the implementation research literature on community-based interventions to improve postpartum contraception use and may inform other CHW programs in similar contexts.

## Background

Contraceptive access is an important precursor that supports women and girls in making decisions pertaining to their sexual and reproductive health (1–4). The postpartum period is a particularly critical time to provide women with quality contraceptive counseling and services. Since short interval pregnancies are associated with adverse maternal and child outcomes, the World Health Organization (WHO) recommends at least 24 months of spacing between births (5–7). However, the unmet need for postpartum contraception remains high globally (7–9) despite postpartum family planning being an important strategy to address maternal mortality in low and middle-income countries (LMICs) (10).

In Nepal, 21% of married women have an unmet need for contraception (11) with likely higher rates among postpartum women (12, 13). Between 1996 and 2006, modern contraceptive use among married women increased from 26 to 44%, and has remained unchanged since (11, 14). Forty-two percent of women in Nepal do not attend any postnatal visit, an important missed opportunity for counseling, initiation, and continuation of voluntary postpartum contraception (15). Only an estimated 13% of women who had live births reported being counseled on family planning after delivery (16). Contraceptive counseling and access are especially challenging in Nepal’s remote, hilly areas where the nearest health facility can be hours away on foot, driving a disproportionately higher unmet need for contraception (17, 18).

Nepal’s cadre of Female Community Health Volunteers (FCHVs) has historically played a significant role overcoming barriers to maternal and child care (19–21). Such community health workers (CHWs), who typically belong to the communities they serve, can help address social and cultural barriers and expand access through home-visits, thereby connecting communities with facility-based resources (22). FCHVs, however, as “volunteers” have faced issues with quality of care and inadequate supervision structures (19). The Government of Nepal endeavors to design, implement, and evaluate improved CHW care models in pursuit of universal health care (23). In recent years, increasing evidence advocates for CHWs to receive training, salaries, and substantial support to optimize their role in achieving universal health coverage (24). There is limited evidence, however, assessing the impact of such professionalized CHW models on reproductive, maternal, newborn, and child health (RMNCH) outcomes in Nepal at scale and over time.

To address this evidence gap, *Nyaya Health Nepal*, a Nepali non-governmental organization and *Possible*, a US-based non-profit organization, partnered with the Government of Nepal Ministry of Health and Population to design, implement, and evaluate a pilot integrated RMNCH intervention (25). The intervention, delivered by CHWs, sought to reach an estimated population of 300,000 in two rural districts. In line with WHO recommendations (24), CHWs were local, full-time, salaried, trained and supervised employees, and used a mobile platform that facilitated counseling support and longitudinal data collection to deliver the intervention (25, 26). Contraceptive counseling was integrated into this bundled intervention based on evidence supporting the effectiveness of integrated contraception interventions over stand-alone ones (8, 27, 28). In a previously published analysis of a sub-population receiving this intervention, we found that postpartum women had twice the odds of using modern contraception one-year post-intervention compared to pre-intervention (29).

We sought to study both the implementation process and effectiveness of this integrated RMNCH intervention using a type 2 hybrid effectiveness-implementation study design (30, 31). Guided by the RE-AIM framework, we conducted a non-randomized, single arm, pre-post study with repeated measurements, and nested qualitative methods to assess the intervention’s *reach* (did the target population receive the intervention?), *effectiveness* (was the intervention effective?), *adoption* (were relevant stakeholders willing and able to initiate it?), *implementation* (was the intervention delivered as planned?), and *maintenance* (can the intervention be sustained?) (31, 32). We have previously reported the bundled intervention’s outcomes on antenatal and postnatal care (26). In this paper, we focus on postpartum contraceptive outcomes associated with the integrated RMNCH intervention over a five-year period in two districts.

## Methods

### Study Site

We conducted this study in Achham and Dolakha districts in Nepal. Achham, located in the hilly, far western region, suffers from poverty and the aftermath of civil war, with the nation’s highest neonatal and under-five mortality rates at 41 per 1000 live births and 69 per 1000 live births, respectively (16, 33). Dolakha, located about 6 hours by road from the capital Kathmandu, suffered a major earthquake in 2015 which devastated its infrastructure. Despite their locations in different regions in Nepal, Achham and Dolakha share similar challenges to healthcare access due to hilly terrain, long distances to health care facilities, and poor road infrastructure.

During the study period, *Possible/Nyaya Health Nepal* managed a government hospital in each district through a public private partnership model. In addition to these two hospitals, each ward (the smallest administrative unit) had a health post that provided basic primary care services. Larger primary health care centers staffed by a medical officer or physician existed every few wards. Travel to health facilities could take up to five hours and be significantly lengthened during the monsoon season. Modern contraceptive availability varied by type of method in both districts; condoms, pills, and injectables were relatively consistently available at health posts, primary health care centers, and the two hospitals. Long-acting methods, including IUDs and implants, were available at a limited number of facilities where there were trained staff. Sterilization procedures were only performed at the hospitals.

We implemented the RMNCH intervention in stepped fashion in both districts between 2015 and 2019, with 11 non-random steps in implementation expansion that we refer to as “hubs”. Each “hub” was a cluster of 2 to 14 wards. Intervention expansion was determined by local stakeholders, and was subject to real-world implementation constraints including unanticipated delays. In 2017, following Nepal’s constitutional transition to federalism, the intervention catchment areas were restructured into municipalities. Since “hubs” were defined prior to this restructuring, they do not completely align with current municipal boundaries.

### Study Design

Guided by the RE-AIM framework, we employed a non-randomized, single arm, pre-post study design with repeated measurements to assess the outcomes in Table 1, and used nested qualitative data collection to further contextualize our results. Implementation of the RMNCH intervention could not be randomized as it was pragmatic and stepped, with roll-out sequence determined by local needs and constraints. Further, data collection was built into CHWs’ regular care delivery to participants, rendering it practically infeasible and unethical to include comparison sites without providing healthcare services. For this analysis, we calculated postpartum modern contraceptive outcomes pre– and post-implementing the bundled intervention, with repeated measures.

**Table 1:** Reach, Effectiveness, Adoption, Implementation and Maintenance (RE-AIM) metrics for postpartum contraception through bundled RMNCH intervention.

| RE-AIM Construct | Metric |
| --- | --- |
| Reach | 1. Proportion of women with live births who were offered and received balanced counseling at least once within their first year postpartum<br>2. Proportion of married women of reproductive age who were offered and received balanced counseling at least once during each post-intervention year |
| Effectiveness | Primary outcome: Postpartum modern contraceptive prevalence rate (PPCPR) for women, stratified by early postpartum (0-5 months) and late postpartum (6-11 months) periods<br>Secondary outcomes:<br>1. Contraceptive method mix, stratified by early postpartum (0-5 months) and late postpartum (6-11 months) periods<br>2. Proportion of postpartum women using lactational amenorrhea who switched to another modern contraception method<br>3. Proportion of infants (0-5 months) who were exclusively breastfed |
| Adoption | Time from participant enrollment to implementation of postpartum BCS within each intervention step (hub) |
| Implementation | Fidelity to workflow, measured as timing of first BCS session (median days postpartum when first BCS session was offered) |
| Maintenance | No quantified metric; qualitative discussion |

### Study Population

All married women of reproductive age (“MWRA”; 15-49 years) and children under two years old living in the study sites were eligible to receive the integrated RMNCH intervention. The intervention focused on MWRA based on guidance from local stakeholders on cultural norms. We employed a census approach, with CHWs attempting to visit all homes in the study sites and offering enrollment to the integrated RMNCH care program for all eligible persons (34). Individuals were excluded from the study if they declined enrollment in the intervention or enrolled in the CHW program without consenting to data use for research purposes.

Study participants for qualitative data included a subset of enrolled women, and the program team that implemented the intervention, including community health nurses (who supervised CHWs), CHWs, and program managers. Program team members were purposively selected considering the duration of their engagement in the program and representation from different intervention sites. Among intervention participants, we purposively selected from those who had given birth during the study period to glean insights from their experiences of engaging with CHWs from pregnancy through postpartum.

### Integrated RMNCH intervention

The integrated RMNCH intervention included five components: (i) home-based antenatal and postnatal care counseling and care coordination, (ii) continuous and active pregnancy screening for all reproductive-age women, (iii) Community-Based Integrated Management of Newborn and Childhood Illness for children under age two, (iv) group antenatal and postnatal care, and (v) person-centered contraceptive counseling using the *Balanced Counseling Strategy* (BCS). CHWs delivered this bundled intervention using CommCare, an open-source, mobile platform that guided counseling, provided decision support, and allowed for offline data collection. The full details of the intervention and its implementation, and sociodemographic characteristics of the study site are published elsewhere (25, 26, 29).

CHWs received training on CommCare use, data collection, informed consent, and clinical topics. For the contraceptive counseling component, training materials emphasized best practices including shared decision-making, respect for autonomy, and anticipatory guidance on potential side effects (29, 35–37). CHWs typically served one ward (population: 1000 to 6000), and larger wards had two assigned CHWs. CHWs visited all enrolled MWRA every three months to actively screen them for pregnancy using at-home urine pregnancy tests. Women who reported undesired pregnancies were referred for options counseling and abortion care. Pregnant women with desired pregnancies received monthly home visits where CHWs provided gestational age-specific counseling and screened for pregnancy danger signs. Pregnant women also received four group antenatal care sessions at local health posts that were co-facilitated by CHWs and government nurse midwives. CHWs also visited women after they gave birth and visited all children monthly until two years of age for counseling, and screening and referrals for early childhood illnesses.

### The contraceptive counseling intervention component

General counseling on available contraceptive options was integrated into the eighth month antenatal home visit and the group antenatal care sessions for pregnant women. For postpartum women, CHWs offered interactive, person-centered contraceptive counseling using an adapted *Balanced Counseling Strategy* (BCS) during home visits. BCS is a toolkit to facilitate high-quality, individualized contraceptive counseling (38–40). It uses a series of questions to narrow method choices to those most consistent with a person’s reproductive values and preferences. The counseling is guided using visual aids that illustrate the relative effectiveness of contraceptive methods. In Nepal, BCS has been used in facility-based post-abortion care settings to improve contraceptive method mix (41).

We adapted BCS for CHW-delivered postpartum contraceptive counseling, and built counseling modules into CommCare to guide counseling sessions. We also adapted BCS to include materials on lactational amenorrhea (LAM), and given cultural norms, trained CHWs to involve husbands and mothers-in-law. CHWs were also trained to offer repeat counseling sessions during the first year postpartum, if desired by the participant. Women who selected a contraceptive method through counseling were referred to the nearest health facility where the method was available. CHWs followed up at subsequent home visits to check whether women initiated their chosen methods, help address barriers to access, and provide ongoing counseling regarding potential side effects. CHWs did not distribute contraceptives at home visits.

CHWs also provided counseling on the benefits of breastfeeding and the importance of transitioning from LAM to another effective contraceptive method at six months postpartum, if women did not desire another pregnancy. If women self-reported high-risk medical conditions, such as hypertension, CHWs referred them to the local health post or to the CHW’s community health nurse supervisor, who would visit them to further review safety of contraceptive methods chosen.

During the first year of implementation in the initial two hubs, CHWs visited postnatal women every month until 12 months postpartum and offered BCS at postpartum months 1, 5, and 10. However, CHWs provided feedback that the majority of the women were exclusively breastfeeding at 1 month postpartum and contraceptive counseling at this visit was less relevant. They preferred to focus on screening for high-risk postpartum conditions and counseling on newborn care at the initial monthly visits. In response, we modified the CHW schedule to offer BCS only starting at 3 months postpartum. After the first two months, CHWs visited postpartum women every three months along with all MWRA, and offered BCS during these visits. CHWs continued to visit children monthly up to the age of two years. Although the intervention originally focused on contraception for postpartum women, after the first year of implementation, the program team felt that the person-centered counseling strategy would be beneficial to all women considering contraception. This led to an expansion of BCS to *all* enrolled married women of reproductive age in the study sites regardless of postpartum status. By design, women were not offered BCS during situations where they did not need contraception, such as during pregnancy, post-menopause, or if they had hysterectomies or sterilizations.

### Implementation strategies

We employed several implementation strategies to optimize the intervention. These included training for CHWs, and their longitudinal engagement with intervention recipients through regular home visits. CHWs participated in regular meetings and received close, ongoing supervision from community health nurses, who helped plan and oversee their tasks and problem-solve any challenges. Further, the CommCare tool provided a standardized workflow and algorithm for care-delivery. The overall implementation process was iterative, and adjustments were made to the workflow based on feedback from those implementing it. Further, as described above, we adapted intervention components to fit local needs and norms.

### Data collection

CHWs collected data on contraceptive use through CommCare, except during pre-intervention data in the oldest hub where they used SurveyCTO, a different mobile platform. In addition to built-in data validation mechanisms within CommCare, the research team regularly conducted data quality checks during data collection. Although we originally planned for data collection through December 2020, Nepal underwent a national lockdown because of the COVID-19 pandemic at the end of March 2020. Given this constraint, our analysis includes hubs that had both: i) pre-intervention data, and ii) at least one year of post-implementation data before April 2020. Based on the timing of intervention roll out, 7 of the total 11 hubs had at least one full year of post-implementation data before the lockdown, so our analysis spans these 7 hubs (50 wards) across Achham and Dolakha districts. For all seven included hubs, we used data available through December 31, 2020.

The time from enrollment to complete roll-out of all RMNCH intervention components *within* a hub varied, ranging from 1 to 10 months, mostly due to differing logistical constraints. As noted previously, the duration *between* expanding the intervention from one hub to another was pragmatic and depended on local government, and varied from 1 to 13 months. At the time we conducted this analysis, the earliest hub had four years of data following the intervention start, and the newer hubs had one year of data each.

During the enrollment phase in each hub, CHWs collected baseline data for all enrolled women. Baseline data included families’ demographic characteristics and a birth history for the preceding two years. CHWs also asked about baseline contraceptive use and type, which served as the pre-intervention data for our main effectiveness outcome (PPCPR). Following enrollment, as CHWs delivered the intervention, they prospectively recorded data on postpartum contraceptive use and balanced counseling during postnatal home visits or pregnancy screening visits. CHWs recorded data on exclusive breastfeeding during follow-up visits with children up to six months of age. These data on contraception and breastfeeding served as our post-intervention data.

*Qualitative data:* A female qualitative researcher (co-author R. Khatri), who was not a part of the care delivery team, used an ethnographic approach including interviews, focus group discussions (FGDs), and participant observation to better understand the implementation process. RK conducted 17 semi-structured individual interviews in Nepali with intervention recipients, community health nurses, and program managers, and three FGDs each with 10-11 CHWs. For participant observation, she accompanied CHWs during home visits and maintained regular field notes. All interviews were audio recorded, transcribed, and translated into English by a professional transcription service.

### Data analysis

#### Quantitative data

*Reach:* We used data from CHW home visits to assess the *reach* of postpartum contraceptive counseling using BCS in each hub. We first determined one-year observation periods following intervention roll-out for each hub (Figure 1). For example, in one hub, postpartum contraceptive counseling component started in February 2017, so we defined the first post-intervention year as February 2017 – January 2018, and so on. Thus, given the stepped implementation, the post-intervention periods for each hub corresponded to its intervention roll-out timing. We describe the observation periods for each hub in a supplementary table. Within each hub, we identified all women who had a live birth during each time period, and calculated the proportions who had at least one CHW visit where BCS was offered, completed, and a contraceptive method was chosen within their first year postpartum. Similarly, we calculated the proportion of ***all*** married women of reproductive age who were offered and received balanced counseling at least once during each post-intervention year. We excluded CHW visits where women were pregnant, postmenopausal and/or had sterilization or hysterectomies because they were not offered contraceptive counseling.

**Figure 1:**
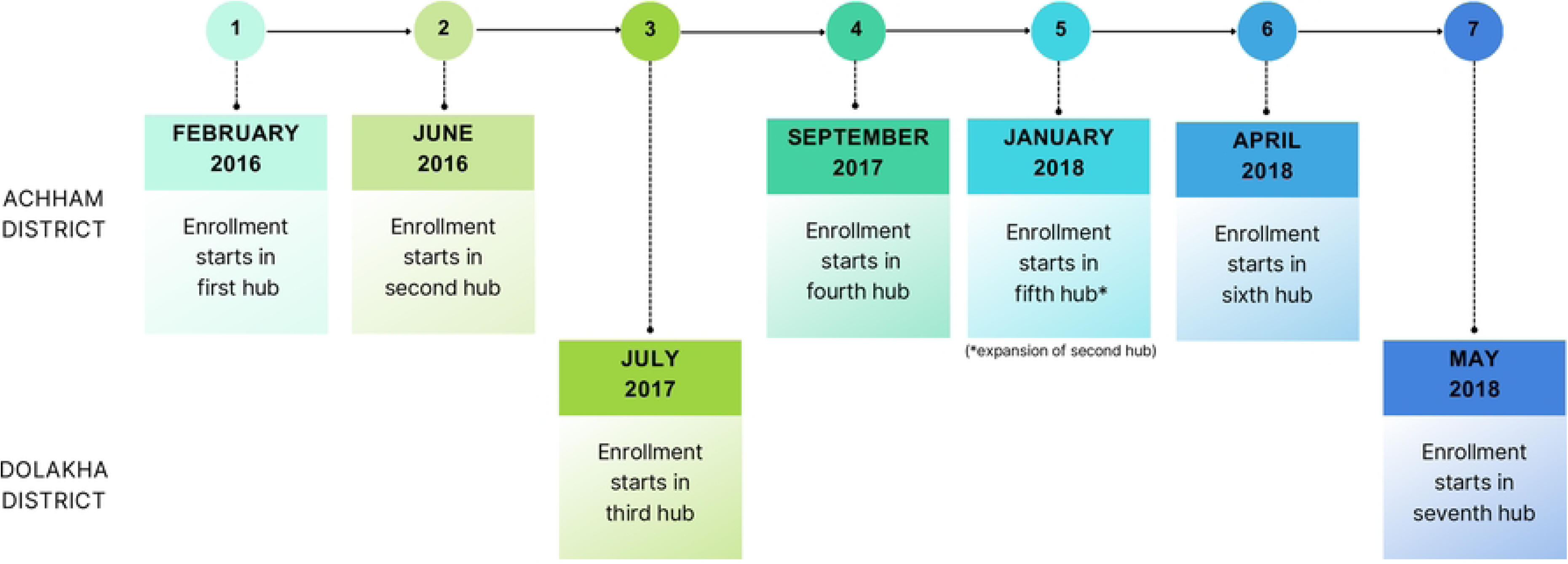
Stepped implementation timeline (not to scale)

*Effectiveness*: We calculated the main *effectiveness* outcome (PPCPR) at the ward level, disaggregated by 0-5 months and 6-11 months postpartum periods. For pre-intervention data in all hubs, we summarized the proportion of postpartum women in each ward who reported using a modern method of contraception at enrollment. In one hub (comprising eight wards), due to a change in workflow, postpartum women were not asked about contraception during the enrollment period. We calculated a proxy pre-intervention PPCPR for this hub, using the first available, non-missing contraception data recorded in the four months immediately following the enrollment period. To calculate post-intervention PPCPR for all hubs, we chose the last six months of each time period for all wards, during which we identified the most recent record of contraceptive use for each woman, also stratified by 0-5 or 6-11 months postpartum periods. We repeated this calculation for each year post-intervention with data available and also used these data to summarize contraceptive method mix.

We summarized the ward level, unadjusted postpartum contraceptive prevalence rates over intervention periods (years). We fitted a mixed-effects Poisson regression model to assess changes in ward-level PPCPR post-intervention compared to pre-intervention, stratified by early (0-5 months) and late (6-11 months) postpartum periods. We used a binary variable *intervention* as the independent variable, representing pre-intervention (0) or post-intervention (1). We adjusted for random effects of *hub*, assuming intervention implementation variations between hubs. We also included *district (Achham* or *Dolakha)* to account for all measured and unmeasured differences across both implementing contexts. Our models thus assessed the association between *intervention* and PPCPR, adjusting for district, and random effects of hubs. In a second set of models (also stratified by early and late postpartum periods), we used a discrete *time period* variable representing the number of years of intervention implementation (0 representing pre-intervention, or intervention years 1, 2, 3, and 4) instead of the binary “intervention” variable to assess whether PPCPR was associated with duration of intervention. We exponentiated coefficients from all models to obtain rate ratios.

The following metrics were calculated using *only* post-intervention programmatic data:

*Secondary effectiveness variables:* To calculate the proportion of women who switched to a modern contraception method after LAM, we identified the subset of women who reported using LAM at any point within their first six months postpartum. Among these individuals, we identified the first non-missing, non-LAM contraceptive use within their first year postpartum. If no contraception data were recorded during 6-11 months postpartum for those previously on LAM, we assumed they did not switch to a modern method, even if they did so after 365 days. We calculated exclusive breastfeeding rates for each observation period using the last available exclusive breastfeeding status for each child under six months old.

*Adoption*: We summarized time (in months) from enrollment to the start of postpartum contraceptive counseling using BCS in each hub from a project Gantt chart that was updated throughout the implementation period.

*Implementation:* We calculated the median days postpartum when women were first offered BCS in each hub for each available time period to assess fidelity to the intervention workflow. In hubs 1 and 2, BCS should have been offered within the first month or first CHW visit postpartum. After updating the workflow (as described above) based on CHW input, this changed to offering BCS at the first contact or starting at 3 months postpartum across all hubs.

We conducted all quantitative analyses using SAS 9.4 and used PowerBI and Microsoft Office to generate graphics. We used R to fit the mixed effects Poisson regression model.

#### Qualitative data

For qualitative data analysis, all transcripts were uploaded into Dedoose Version 9.0.17. Author R. Khatri developed codes using an inductive content analysis approach to qualitatively understand how the integrated intervention influenced postpartum contraceptive uptake, including facilitators and barriers to CHW provision of care and experiences of the participants enrolled in the intervention. The codes also included new concepts introduced by the data. We extracted themes and subthemes relevant to the implementation of contraceptive counseling and postpartum contraception utilization.

#### Ethics approval

The Nepal Health Research Council (461/2016), the Brigham and Women’s Hospital institutional review board (2017P000709/PHS), and Mount Sinai institutional review board (MSSM IRB-18-01091) provided human subjects approval for the study. The Boston Medical Center (H-38196) institutional review board exempted the study since the first author W. Wu had moved from Brigham and Women’s Hospital to Boston Medical Center after the data collection process was already complete. At that point, only de-identified data were being used for analysis and manuscript preparation. Since the study already had approval from the Nepal Health Research Council and Brigham and Women’s institutional review boards, the primary author’s scope of work was not considered human subjects research that the Boston Medical Center was engaged in and was considered exempt.

During enrollment into the RMNCH intervention, CHWs read the consent form and obtained verbal informed consent to enroll households and individuals into the CHW program, provide care, and use their programmatic data for research. Consent was documented electronically in CommCare. Households and individuals could choose to participate only in the care program and decline use of their data in research activities. Their data were excluded from this study. For the children who were under two years, their mothers provided verbal consent for enrollment into the care program and use of their data for research. For the qualitative interview and FGD participants, the research team member read a structured script and obtained verbal informed consent. Consent included use of de-identified quotes for publication purposes. We opted for verbal consent since illiteracy was a concern in our study population, which made it infeasible and unethical to ask for written consent. Further, in the local context of our study sites, fingerprinting on paper had election/political connotations, and participants would have been hesitant to participate if asked to provide a fingerprint. We also took into consideration the local rules and local IRB at the time, which did not require written consent. Finally, given the study’s low level of risk, i.e. consent to use of data from routine clinical care, verbal consent was considered appropriate.

## Results

### Reach

We found that most (83%-97%) women who had live births in each hub were offered BCS at least once in their first year postpartum (Table 2). Agreement to participate and actual completion of counseling sessions was lower, and ranged from 39% to 81% across the hubs. Among those who completed at least one session, nearly all chose a contraceptive method at the end of the session.

**Table 2:** Balanced counseling for postpartum women (who had live births)

| District | Hub | Postpartum women<br>n (%) | Intervention year (hub-specific) |  |  |  |
| --- | --- | --- | --- | --- | --- | --- |
|  |  |  | Year 1 | Year 2 | Year 3 | Year 4 |
| Achham | 1 | *Participated in CHW visit* | - | 542 | 536 | 527 |
|  |  | *Was offered balanced counseling* | - | 530 (83%) | 508 (91%) | 509 (93%) |
|  |  | *Completed balanced counseling* | - | 373 (59%) | 361 (64%) | 300 (55%) |
|  | 2 | *Participated in CHW visit* | 454 | 441 | 389 | - |
|  |  | *Was offered balanced counseling* | 449 (92%) | 428 (94%) | 380 (96%) | - |
|  |  | *Completed balanced counseling* | 318 (65%) | 260 (57%) | 153 (39%) | - |
|  | 4 | *Participated in CHW visit* | 458 | 425 | - | - |
|  |  | *Was offered balanced counseling* | 443 (90%) | 408 (89%) | - | - |
|  |  | *Completed balanced counseling* | 397 (81%) | 359 (78%) | - | - |
|  | 5 | *Participated in CHW visit* | 129 (96%) | 122 (98%) | - | - |
|  |  | *Was offered balanced counseling* | 125 (93%) | 121 (97%) | - | - |
|  |  | *Completed balanced counseling* | 82 (61%) | 41 (33%) | - | - |
|  | 6 | *Participated in CHW visit* | 543 | - | - | - |
|  |  | *Was offered balanced counseling* | 511 (92%) | - | - | - |
|  |  | *Completed balanced counseling* | 402 (73%) | - | - | - |
| **Dolakha** | 3 | *Participated in CHW visit* | 260 | 298 | - | - |
|  |  | *Was offered balanced counseling* | 256 (97%) | 286 (95%) | - | - |
|  |  | *Completed balanced counseling* | 183 (70%) | 172 (57%) | - | - |
|  | 7 | *Participated in CHW visit* | 252 (99%) | - | - | - |
|  |  | *Was offered balanced counseling* | 247 (97%) | - | - | - |
|  |  | *Completed balanced counseling* | 102 (40%) | - | - | - |

Women’s participation and completion of BCS sessions may be dependent on several factors. Our qualitative data suggest that it was difficult for women to spare time for counseling, especially during seasons when they had to work in the fields. In addition, spouses or other family members could also influence their participation as described by a CHW below:

> *I laid out the cards in front of her for counseling and at the same time, her husband arrived. Generally, he is a nice man but I don’t know what happened that day, he got angry. I asked him to stay together for counseling but he didn’t. I was a little scared seeing his face thinking he might say something. His behavior was changed….He told his wife that they have to eat…. His wife was also scared and said that she will use depo [injectable contraception] only….. I collected the cards and kept them in my bag. He later said that they know that they will have a baby after sexual intercourse and they know what to do and I don’t have to tell them.*
>
> FGD with CHWs, Achham

A few other CHWs highlighted women’s lack of autonomy in making decisions around contraceptive use. One CHW described a woman who could not think about using contraceptives without asking her husband, who was working in India at the time. Another CHW described how a woman’s family members were upset that they were not consulted about her contraception choices. The family members scolded the CHW, and asked her to be responsible if anything happened to their daughter-in-law, as described in the quote below. Women’s willingness to accept contraceptive counseling was thus influenced by concerns about their husband’s or family members’ reactions.

> *There is a woman who had used implant. She was fine but when I went near her house, the members of the household scolded her and me….saying I have to be responsible for their daughter-in-law. Her father-in-law was angry that the daughter-in-law didn’t take his/ family’s permission [for contraception use], but consulted with me [CHW]. [They had said] the daughter-in-law would get cancer and her arm would have to be cut off. There was another woman in the neighborhood who was ready to use the implant but because of them, she didn’t use it.*
>
> FGD with CHWs, Achham

According to post-intervention programmatic data, the proportion of all married women of reproductive age who were offered contraceptive counseling using BCS increased over time (Table 3). By the last available post-intervention year for each hub, which roughly corresponded with calendar year 2020, over 95% of MWRA in all hubs had been *offered* BCS at least once. Completion of BCS tended to remain low in most hubs (≤ 20%), except in hub 4 where it was over 50%.

**Table 3:** Balanced counseling for all married, reproductive age women.

| District | Hub | Married, reproductive age women<br>n (%) | Intervention year (hub-specific) |  |  |  |
| --- | --- | --- | --- | --- | --- | --- |
|  |  |  | Year 1 | Year 2 | Year 3 | Year 4 |
| Achham | 1 | Participated in CHW visit | - | 3302 | 4703 | 4252 |
|  |  | Was offered balanced counseling | - | 861 (26%) | 3478 (74%) | 4128 (97%) |
|  |  | Completed balanced counseling | - | 553 (17%) | 1246 (26%) | 1552 (37%) |
|  | 2 | Participated in CHW visit | 2768 | 2651 | 2629 | - |
|  |  | Was offered balanced counseling | 831 (30%) | 2284 (86%) | 2605 (99%) | - |
|  |  | Completed balanced counseling | 612 (22%) | 351 (13%) | 488 (19%) | - |
|  | 4 | Participated in CHW visit | 2749 | 2546 | - | - |
|  |  | Was offered balanced counseling | 2076 (76%) | 2497 (98%) | - | - |
|  |  | Completed balanced counseling | 1525 (55%) | 1571 (62%) | - | - |
|  | 5 | Participated in CHW visit | 798 | 705 (89%) | - | - |
|  |  | Was offered balanced counseling | 705 (88%) | 695 (99%) | - | - |
|  |  | Completed balanced counseling | 252 (32%) | 138 (20%) | - | - |
|  | 6 | Participated in CHW visit | 3842 | - | - | - |
|  |  | Was offered balanced counseling | 3691 (96%) | - | - | - |
|  |  | Completed balanced counseling | 2455 (64%) | - | - | - |
| Dolakha | 3 | Participated in CHW visit | 2928 | 2673 | - | - |
|  |  | Was offered balanced counseling | 2022 (69%) | 2650 (99%) | - | - |
|  |  | Completed balanced counseling | 290 (10%) | 509 (19%) | - | - |
|  | 7 | Participated in CHW visit | 1842 | - | - | - |
|  |  | Was offered balanced counseling | 1791 (97%) | - | - | - |
|  |  | Completed balanced counseling | 362 (20%) | - | - | - |

### Effectiveness

The box plots of unadjusted ward-level postpartum contraceptive prevalence rates by intervention time period (Figure 2), suggest an association between the intervention and PPCPR in both the 0-5 and 6-11 months postpartum women populations.

**Figure 2:**
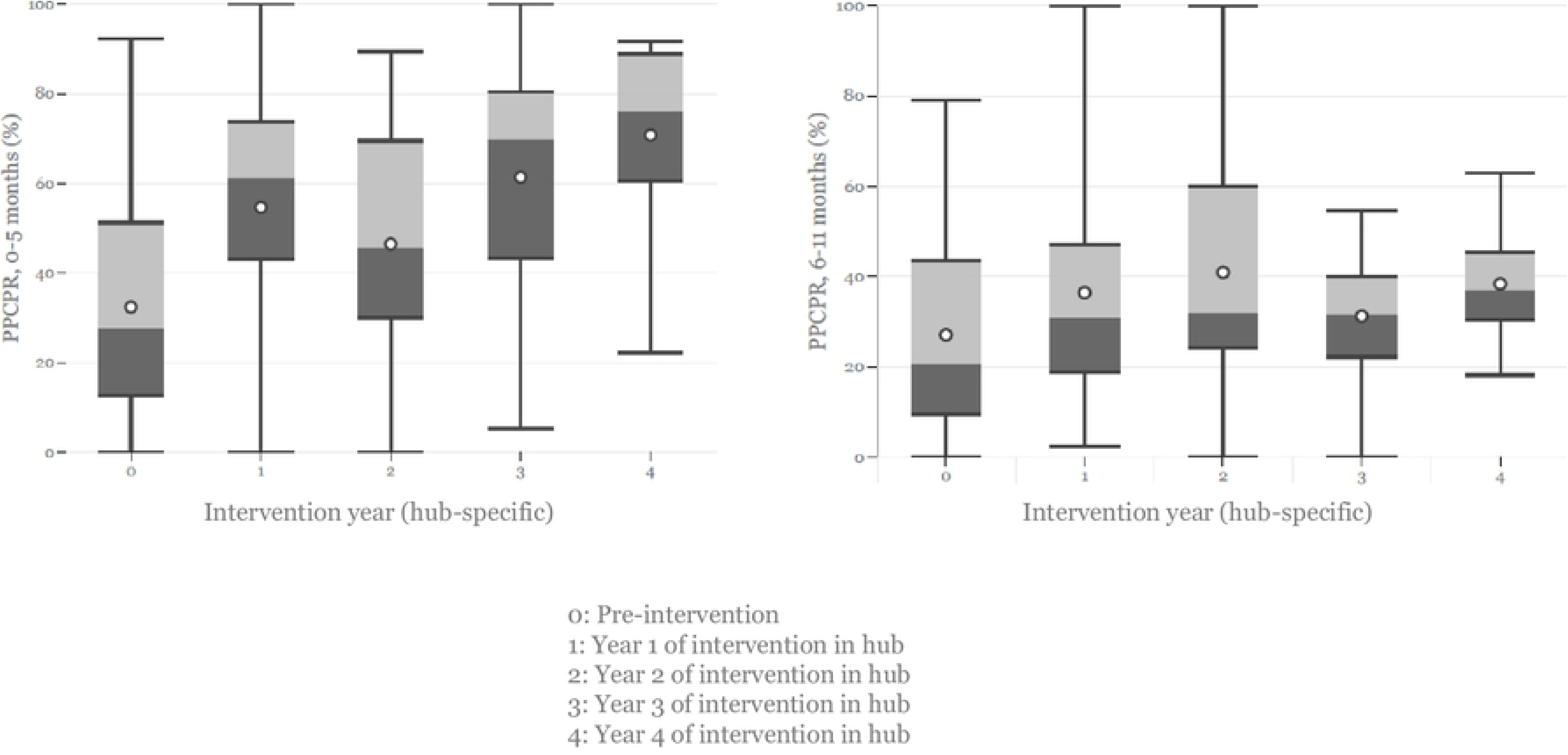
Unadjusted ward-level postpartum contraceptive prevalence rates by intervention time period.

Adjusting for district and random effects of hubs, the ward-level PPCPR for women between 0-5 months postpartum was higher post-intervention compared to pre-intervention (Rate ratio: 2.20; 95% CI: 1.96, 2.48). Similarly, adjusting for district, and random effects of hubs, the ward-level PPCPR for women between 6-11 months postpartum was higher post intervention compared to pre-intervention (RR: 1.70; 95% CI: 1.50, 1.93).

There was also an association between PPCPR in both early and late postpartum periods and the number of years of the bundled intervention (Table 4b). In year 4 (which we only observed in hub 1), the ward-level PPCPR for women between 0-5 months postpartum was 3.16 times the pre-intervention rate (95% CI: 2.63, 3.79). The ward-level PPCPR in Dolakha also tended to be higher compared to that in Achham, especially in the late postpartum period (Tables 4a and 4b).

**Table 4a:** Associations between RMNCH intervention and ward-level postpartum contraceptive prevalence rate.

|  | Postpartum contraception prevalence rate |  |  |  |
| --- | --- | --- | --- | --- |
|  | Early postpartum (0-5 months) |  | Late postpartum (6-11 months) |  |
|  | Rate Ratio (95% CI) | p-value | Rate Ratio (95% CI) | p-value |
| **Pre-intervention (Year 0)** | 1.00 |  | 1.00 |  |
| **Post-intervention** | 2.20 (1.96, 2.48) | <0.001* | 1.70 (1.50, 1.93) | <0.001* |
| **District - Achham** | 1.00 |  | 1.00 |  |
| **District - Dolakha** | 1.33 (1.06, 1.69) | 0.006* | 2.60 (1.69, 3.99) | <0.001* |

**Table 4b:** Associations between years of RMNCH intervention and ward-level postpartum contraceptive prevalence rate.

|  | Postpartum contraception prevalence rate |  |  |  |
| --- | --- | --- | --- | --- |
|  | Early postpartum (0-5 months) |  | Late postpartum (6-11 months) |  |
|  | Rate Ratio (95% CI) | p-value | Rate Ratio (95% CI) | p-value |
| **Pre-intervention (Year 0)** | 1.00 |  | 1.00 |  |
| **Year 1** | 2.17 (1.91, 2.47) | <0.001* | 1.51 (1.31, 1.74) | <0.001* |
| **Year 2** | 1.93 (1.67, 2.23) | <0.001* | 1.80 (1.55, 2.11) | <0.001* |
| **Year 3** | 2.50 (2.09, 2.97) | <0.001* | 2.01 (1.64, 2.46) | <0.001* |
| **Year 4** | 3.16 (2.63, 3.79) | <0.001* | 2.24 (1.77, 2.82) | <0.001* |
| **District - Achham** | 1.00 |  | 1.00 |  |
| **District - Dolakha** | 1.4 (1.15, 1.69) | <0.001* | 2.76 (1.9, 3.98) | <0.001* |

### Other effectiveness outcomes

Contraceptive method mix (Figure 3) for women 0-5 months postpartum showed an overall increase in LAM use from pre-intervention to post-intervention. Injectables were the second most common method of contraception in the 0-5 month postpartum period, and the most common method in the 6-11 month postpartum period. Injectable use was particularly high in Dolakha district, with up to 57% of 6-11 month postpartum women using injectables by the second post-intervention year in hub 3. There also appeared to be an increase in long acting and permanent methods (LAPM) in the 6-11 month postpartum period in Achham district, and an unanticipated increase in traditional methods in hub 7 post-intervention.

**Figure 3:**
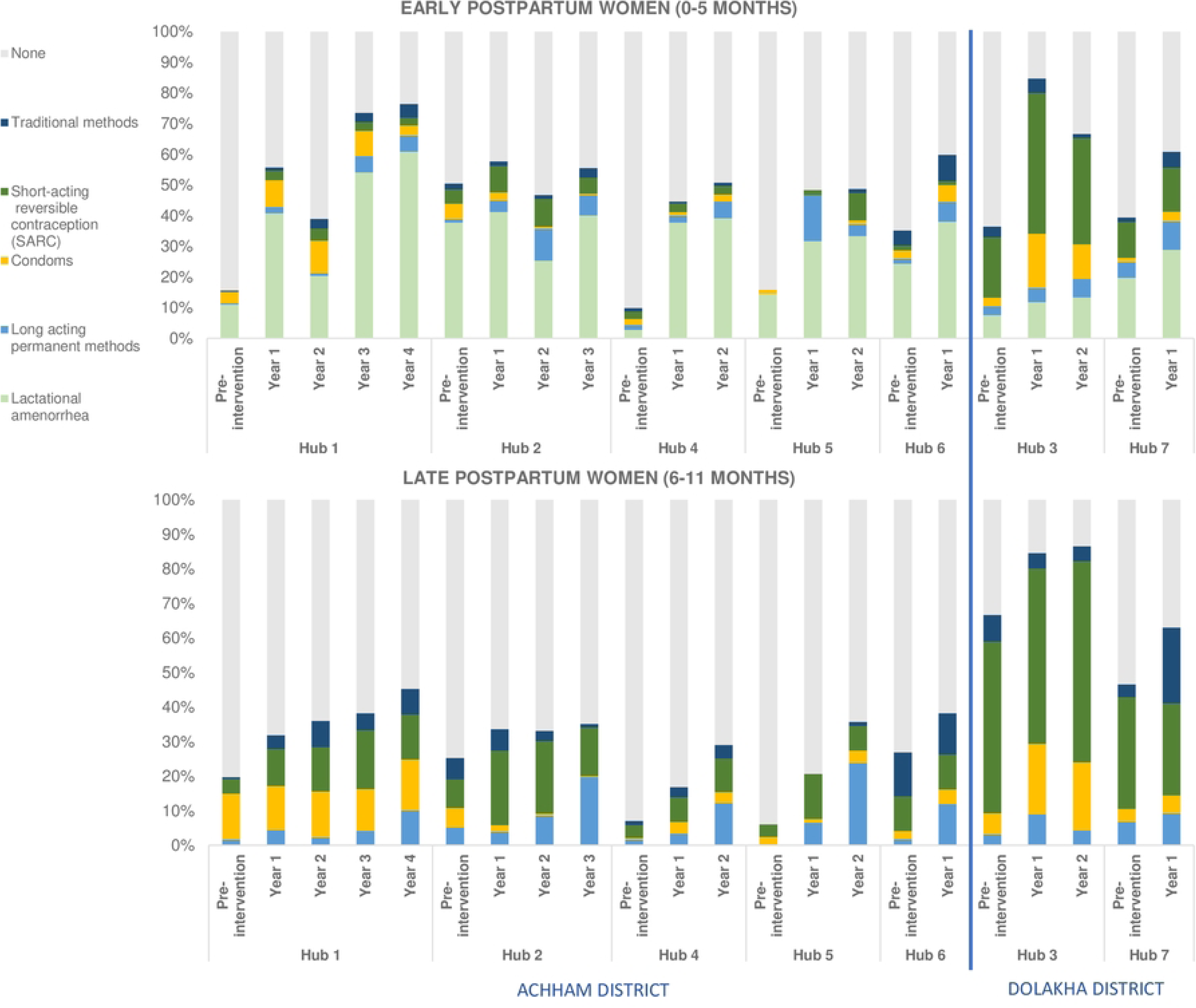
Postpartum contraceptive method mix.

The proportion of women on LAM within 0-5 months postpartum who switched to another modern method of contraception remained quite low, and ranged from 10% to 30% in most hubs, except for hub 3 where it was over 60%. Exclusive breastfeeding among infants (0-5 months old) was quite high (Figure 4) in all hubs and appeared to increase over time post-intervention.

**Figure 4:**
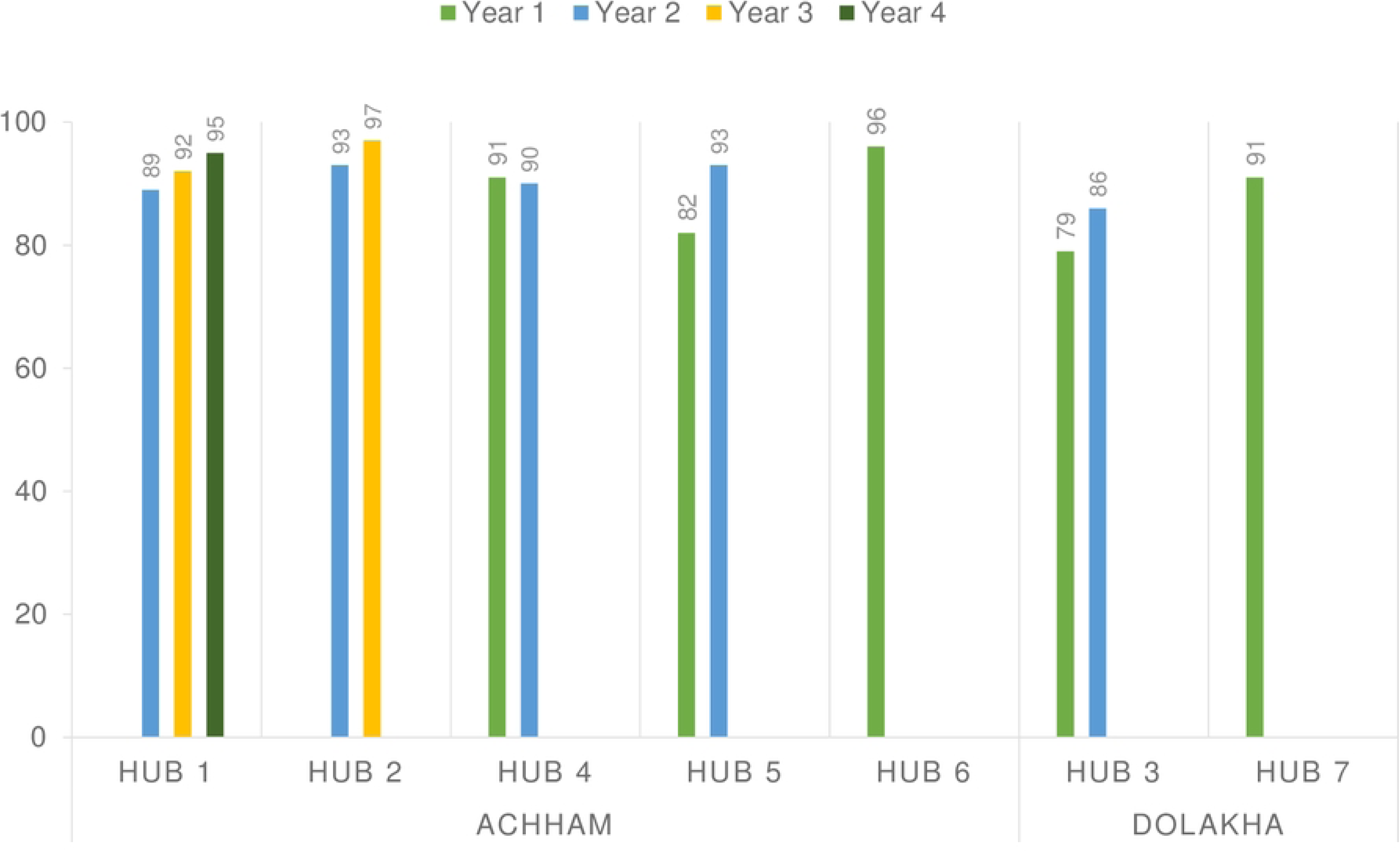
Proportion of infants (0-5 months) who were exclusively breastfed (%)

One likely determinant for intervention effectiveness was the trusting longitudinal relationships the CHWs built with women in their community. The overall trust women had in the CHWs likely contributed to the apparent association of the intervention with increase in contraceptive use postpartum.

> *They share with us, considering us very close to them; even things they won’t share with their sisters-in-law. They ask us what to do in certain situations. Even though we are relatively young, women from our mothers’ age group also share things without any hesitation. When they do so, we feel that they trust us*.
>
> *Initially, they hesitated to answer and wouldn’t open up when asked about family planning but now they treat us as their friends and openly tell us. Some of them will not tell others in the village about their contraceptive use but will tell us what they are using*.
>
> *We go everyday and if some problems happen, we go there as well. We have understood them and their problems. They trust us because we have been able to address their issues*.
>
> FGD with CHWs, Achham

Participants from both Dolakha and Achham districts also appreciated the benefits of CHWs visiting them to discuss counseling for contraceptives.

> *Who understands the plight of women? Earlier, our life was like that of cattle…got married, gave birth but we knew nothing. Now that they [CHWs] come to our doorsteps to teach us these things, we can’t explain how happy we are.*
>
> Intervention participant, Dolakha

> *She teaches us things we didn’t know, new things. She explains things we don’t understand. It has made things easy [to learn].*
>
> Intervention participant, Achham

In addition to building trust among women in the community, CHWs also noted improving relationships with facility-based staff as they implemented the intervention. CHWs demonstrated the capacity to help bridge care from the home to the health facility.

> *People in the community call me and trust me when they are facing any troubles; in those times I feel like I have been able to do something for the community. People in the health post also remember us. They tell us about women who didn’t come for visits and ask us to remind them. At this time, we feel recognized for our work.*
>
> FGD with CHWs, Achham

Barriers that the CHWs noted for contraceptive utilization included negative perceptions and fear of side effects. For instance, one CHW observed:

> *To make them understand about family planning has been very difficult because they think use of family planning will cause cancer, and other problems.*
>
> FGD with CHWs, Achham

During a FGD, CHWs shared that some women expressed a fear of infertility after using contraceptives because of some instances where women reported difficulty conceiving again after using some contraceptives like injectables:

> *They think that using family planning measures early on causes infertility. We had gone to meet [a postpartum woman] who had two days left to reach 45 days. We did the counseling and she went the next day after completing 45 days and used depo [injectables]. Now, her child is 3 years old. She hasn’t had another baby; she had discontinued depo; her husband is also at home. Now, they are not able to have another baby; she even went to [the hospital] for treatment. She told me that I made her use that injection and now she is not able to give birth to a son.*
>
> FGD with CHWs, Achham

One CHW noted that some women did not feel secure with having only one son and wanted more sons; therefore, they do not want to use contraceptives. Son preference seems to be one underlying reason for fear around contraceptive use. Another CHW reported that women feared permanent sterilization would weaken their physical strength and hinder their ability to perform farm work. CHWs reported a similar fear among women regarding vasectomies:

> *We have not been successful in permanent measures because if we tell the woman about a permanent family planning camp, she will say that she will get weak. They say, “We need to work in such a big field, we have to carry the animals, we will not have anything to eat, we will be weak.” This perception still prevails. We have not been able to get rid of this*.
>
> *Women say that they would do it themselves rather than men…They think men will get weaker. They say that men have to go outside, do hard labor like crushing stones…Women have more burden but they don’t see theirs, rather see that of men*.
>
> FGD with CHWs, Achham

Peer influence was one important factor affecting women’s decisions around contraceptive use. CHWs observed that if somebody in the community had negative experiences with contraception or vice-versa, that would affect others’ uptake of contraception.

> *The effect is both ways. If they have heard that it has done good, then others will say that they will also use it and go about using it.*
>
> FGD with CHWs, Achham

Contraceptive uptake was also affected by access to methods in health facilities. In Dolakha, CHWs noted that women would choose a method during BCS, but were unable to access those methods when they went to the health facilities. A CHW noted:

> *Women from my community who had come for implant and IUCD were given condoms and sent away. A woman shared that she was scolded at the MCH (Maternal Child Health) clinic and therefore, she went to a health post at [hub 7] to keep an implant instead of coming to the […] hospital again.*
>
> CHW, Dolakha

### Adoption

Of the 19 intended municipalities, 10 adopted the RMNCH intervention, and 7 of those adopted it within the anticipated timeline. The intervention’s postpartum BCS component was implemented in all hubs where the bundled intervention was introduced. For all the hubs included in the analysis for this paper, except for one, BCS was implemented as part of postnatal care almost immediately after enrollment was completed. The exception to this was hub 2, where postnatal care was rolled out four months after enrollment completion. Since hub 2 was the second implementation step, this additional time was used to update workflows and data collection tools in the mobile platform based on feedback from the implementation experience in the first hub.

### Implementation

As described above, the intervention used an iterative approach, guided by feedback from the program’s team. After the first year of implementing postpartum contraceptive counseling, the program team proposed a more streamlined home visit schedule whereby postpartum women were visited and offered BCS starting at the third month postpartum, consistent with the general MWRA schedule of home visits every 3 months.

Table 5 indicates the timing in median days postpartum when BCS was offered to women. The post-intervention year 2 for hub 1 and year 1 for hub 2 correspond to the same calendar year (February 2017 – January 2018), during which postpartum women were receiving monthly visits from CHWs. Hence, on average, women were about two weeks postpartum when they were first offered BCS counseling in that year. After the switch in timing of home visits to once every three months postpartum, BCS counseling was offered closer to 90 days postpartum.

**Table 5:**
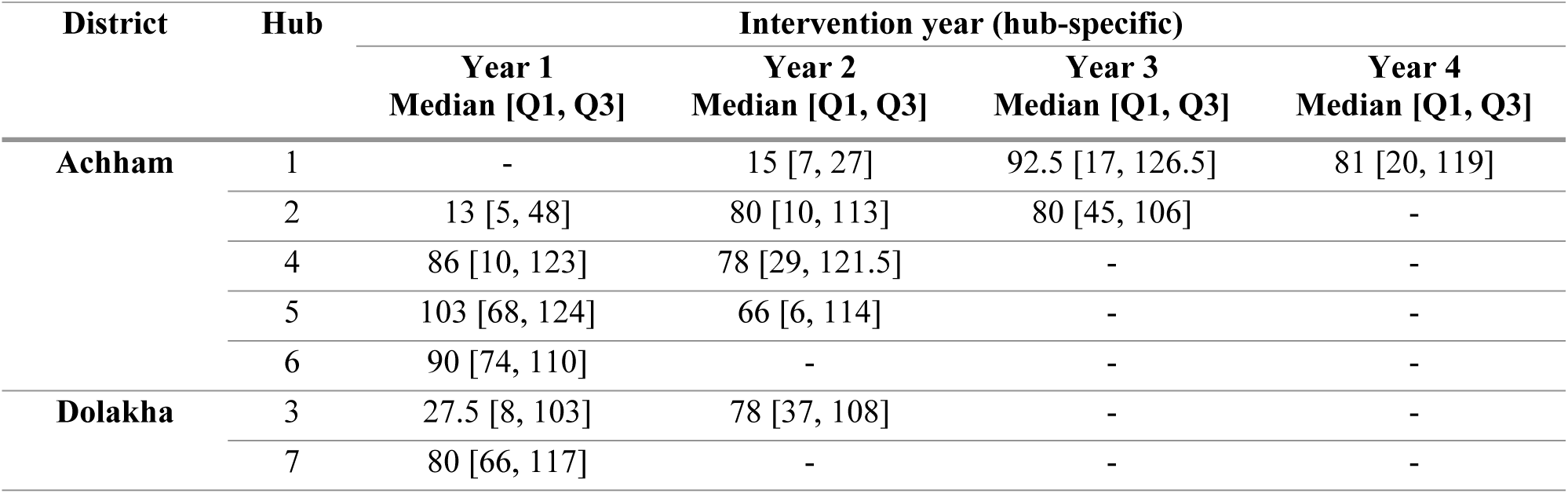
Timing of offering BCS counseling (in days postpartum)

| District | Hub | Intervention year (hub-specific) |  |  |  |
| --- | --- | --- | --- | --- | --- |
|  |  | Year 1<br>Median [Q1, Q3] | Year 2<br>Median [Q1, Q3] | Year 3<br>Median [Q1, Q3] | Year 4<br>Median [Q1, Q3] |
| Achham | 1 | - | 15 [7, 27] | 92.5 [17, 126.5] | 81 [20, 119] |
|  | 2 | 13 [5, 48] | 80 [10, 113] | 80 [45, 106] | - |
|  | 4 | 86 [10, 123] | 78 [29, 121.5] | - | - |
|  | 5 | 103 [68, 124] | 66 [6, 114] | - | - |
|  | 6 | 90 [74, 110] | - | - | - |
| Dolakha | 3 | 27.5 [8, 103] | 78 [37, 108] | - | - |
|  | 7 | 80 [66, 117] | - | - | - |

Additionally, implementation barriers included distance, challenging physical terrains that CHWs traversed to reach some remote communities, and increased travel times during the rainy season. Stable electricity and therefore, mobile phone service and communication with the participants was less reliable during the rainy season. These barriers are also described further in detail elsewhere (26).

> *And the other thing is geographical difficulties. Some areas are so far that we need to stay there…We cannot skip such areas; we have to go there. We have to go to provide services to the people there.*
>
> Supervisor 2, Achham

> *When we work in a community, we face community related challenges like having to walk in the rain, and the public vehicles do not run during the rainy season, our staff have to walk for 6-7 hours…Those kinds of challenges we face in our roles.*
>
> Supervisor 1, Achham

CHWs also brought up social issues like caste as a barrier to delivering care and counseling – some CHWs reported not being permitted in certain spaces because of their caste:

> *People would say that I haven’t washed and dried my hair after menstruation [One has to wash and dry their hair after four days of menstruation to be considered “pure”]. They would tell me to not come saying that I am Dalit [low caste].*
>
> FGD with CHWs, Achham

### Maintenance

BCS continues to be delivered by CHWs in the study sites after the study period. The nonprofit *Nyaya Health Nepal* leads implementation in Achham district, while implementation in Dolakha district has transitioned to the community health program of the Dhulikhel Hospital-Kathmandu University Hospital. Furthermore, the integrated RMNCH intervention described in this study served as the foundation for a similar pilot delivered by community health nurses in two other municipalities in Nepal (42). This new pilot, which also includes BCS, is being led by the Nursing and Social Security Division under the Ministry of Health and Population of the Government of Nepal, with technical assistance from *Possible*.

Our qualitative data explored maintenance of behavior change and the potential for longer term impact from the CHW home visits and counseling. Some women mentioned that they will remember what the CHWs had “taught” them and will try to follow the advice. Most program staff also described believing that the women will follow CHWs’ counseling, despite changes in visit frequency. They anticipate that behavior change built on longitudinal care within an integrated program will have a longer lasting impact rather than standalone one-off programs:

> *We are assured that they will do it because sometimes when we go to their household, we may not meet the person but we will find that the child has been immunized; later on we find out that they have gone for a check-up. For them to continue, this is what gave us hope. I am hopeful, now, that even if we are not in this job, they will continue to do so, they will do it.*
>
> FGD with CHWs, Achham

> *[CHWs] used to tell them things from time to time…And now if CHWs do not visit in a long time, then those things also might be missed. They remember many things such as advantages of family planning, and advantages of visiting the health institution. But some things they might forget in the long run like, about follow-up, right, like these things.*
>
> Supervisor 2, Achham

## Discussion

We studied the impact of a CHW-delivered integrated RMNCH intervention in two districts in rural Nepal on postpartum contraceptive outcomes, and observed an overall increase in modern contraceptive use within one year postpartum. Adjusting for district and hubs, we observed higher post-intervention ward-level PPCPR (0-5 months postpartum) compared to pre-intervention (Rate ratio: 2.20; 95% CI: 1.96, 2.48). Similarly, we observed higher ward-level post intervention PPCPR (6-11 months) compared to pre-intervention (IRR: 1.70; 95% CI: 1.50, 1.93). We observed an overall increase in LAM use with the intervention, and high exclusive breastfeeding rates among infants (0-5 months old) in all hubs, which appeared to increase over time post-intervention. The proportion of women who switched from LAM to another modern contraceptive method tended to be lower, ranging from 10% to 30% in most hubs, except one hub where it was over 60%. Qualitative data helped us to understand factors affecting the effectiveness of the intervention, such as the trust that CHWs developed with women in the community through their longitudinal engagement, and the relationships they built with healthcare providers. CHWs also noted several factors that affected implementation and women’s uptake of contraception, including gender norms, spousal and family involvement, fear of side effects, and the influence of peers.

Our results are consistent with evidence from a review of 35 interventions in LMICs that concluded incorporating contraceptive counseling and services across the reproductive health care continuum is the ideal strategy for improving postpartum contraception (28). CHW programs are an example of how this can be implemented. A quasi-experimental trial in Bangladesh that integrated family planning counseling into an existing maternal and newborn health intervention delivered by CHWs showed that the intervention arm had higher contraceptive prevalence rate at 12 months postpartum, lower short interval pregnancy of <24 months, and lower preterm birth rates (43, 44). Similarly, in our intervention, contraceptive counseling was embedded into a longitudinal CHW program that supports women pre-pregnancy, antenatally and into the extended postnatal period.

Longitudinal follow-up and providing contraceptive counseling into the extended postnatal period is also important as women’s pregnancy intentions and desire to use contraception are dynamic and change over time (45). In Nepal, the most common reason women report for not using contraception postpartum is spousal separation mostly due to migration for work (46). Additional reasons include low self-perceived risk of getting pregnant, fear of side effects, breastfeeding, not having resumed sex postpartum, and amenorrhea (46, 47). Based on DHS data, 25-37% of married women in our study sites lived apart from their spouses, and more than half of those women reported that their spouses are away for more than 12 months (47). Women’s self-perceived risk for pregnancy and contraceptive use are thus influenced by their husbands’ absence, which likely affected their acceptance of contraceptive counseling in our study (48, 49).

We found that nearly all (83%-97%) women across the hubs who had live births were offered BCS counseling at least once in their first year postpartum. These observed rates are quite high, likely due to the clinical decision support built into the CommCare app that prompted CHWs to offer BCS during their visits. However, acceptance and completion of BCS were variable across hubs. In the first year, completion of BCS ranged from 40-81% and in year two, 33-59%. In addition to spousal migration, qualitative data highlighted other factors that influenced whether women accepted contraceptive counseling from CHWs. These included unsupportive family members and husbands, fertility concerns, and limited time to spare for counseling especially during busy farming seasons. Also, as there appeared to be a decrease in BCS completion rates over time in all hubs, which may have been due to intervention participants finding repeated BCS counseling redundant, or diminishing implementation fidelity over time.

Despite increasing rates of facility-based deliveries by skilled birth attendants, about 40% of women in Nepal still do not receive a postnatal check-up and few report receiving family planning counseling after giving birth (47). The quality of family planning counseling that women receive remains concerning, with women reporting poor experiences, including not feeling like they were being listened to (15). BCS embedded within an RMNCH intervention delivered by CHWs offers a community-based, person-centered contraceptive counseling strategy. While this study does not evaluate the quality of contraceptive counseling, in our previous sub-study, women and their husbands found BCS acceptable and noncoercive, and appreciated the visual aids and the description of the range of available methods (29). BCS does have some limitations – while it prioritizes each woman’s reproductive intentions and goals, the counseling algorithm is guided by tiered effectiveness, where the most effective methods are presented first (39).We acknowledge that effectiveness may not be the priority for every woman. As our qualitative data indicate, other social and cultural factors including gender norms, fear of side effects, spousal migration, and opinions of family members may be more influential in decision-making.

Our data shows high rates of exclusive breastfeeding among children under 6 months in the study areas ranging from 79-97%, which is significantly higher than national estimates of 56% (11). This may be related to the intervention’s emphasis on CHW counseling on the benefits of breastfeeding. The median duration of postpartum amenorrhea in Nepal is 6 months (16), suggesting that many women are likely protected from pregnancy in the first 6 months postpartum. We observed high rates of reported LAM use that increased with the intervention years. Our data, however, show relatively low rates of transition from LAM to another effective method of contraception. In settings with early postpartum uptake of short-acting contraceptive methods and high rates of lactational amenorrhea, there is concern that women will discontinue their short-acting method by the time they are no longer protected from pregnancy by LAM (28, 50). Additional research is needed to better understand these low transition rates and how to facilitate the transition from LAM to another effective contraceptive method for women who do not desire pregnancy.

## Limitations

Our study had several limitations. Given the pragmatic study design, we did not have a separate comparison group of hubs, but collected pre– and post-intervention data for each hub. Since all post-intervention quantitative data was programmatic data extracted from the CommCare app as CHWs delivered the intervention, it was neither feasible nor ethical to collect data from other sites where we did not implement the intervention. Further, the implementation process was driven by local stakeholders making it infeasible to employ randomization. Thus, we are limited in our ability to make causal inferences about observed increases in contraceptive uptake despite attempting to account for variations in district and hubs in our analysis.

In addition, we used contraceptive prevalence as the main effectiveness indicator in this study, which is not a person-centered outcome and does not take into account women’s satisfaction with their method (51). Any target prevalence is likely set by public health advocates and not based on individual woman’s decisions and reproductive autonomy (52). Evaluating program success based on contraceptive prevalence may thus inadvertently incentivize providers to prioritize method uptake and lead to coercive practices (52). Future studies may include additional person-centered outcomes including women’s satisfaction with their method, autonomy in making contraceptive decisions, or use of a non-preferred contraceptive method (53).

There were several data-related limitations. For instance, we did not include data on contraceptive counseling with BCS post-abortion and may have slightly underestimated contraceptive counseling among all married women of reproductive age as we did not start collecting this data until the second year of the program. For one hub, we had to calculate a proxy pre-intervention PPCPR using programmatic data because of data limitations, which may have led to overestimation of the pre-intervention contraceptive use. There may have also been some bias introduced since CHWs delivered care and simultaneously collected data. For instance, CHWs may have felt their performance was measured by their success in being able to “convince” women to adopt modern contraception. There are some concerns about reproductive coercion, particularly in hubs with very high contraceptive prevalence. Further qualitative data will need to be collected in these hubs to understand factors that contribute to the high rates.

In addition, since the intervention expansion in all originally planned hubs was delayed for various reasons, we were only able to include data from hubs where we had at least one year of post-intervention data before the national COVID-19 lockdown. Thus, it is possible that the improvements we observed in the hubs included in our analyses may not have been observed in other hubs where the intervention was delayed.

Finally, broader structural and socio-cultural factors that impact access to and use of contraception including poverty, gender inequality, weak supply changes, and poor road infrastructure were not directly addressed through our intervention (17, 34, 54, 55). For instance, while CHWs facilitate community-facility linkages, additional efforts are needed to improve physical access to facilities and quality of care at facilities.

## Conclusion

Despite these limitations, however, this study assesses the real-world implementation of an integrated RMNCH intervention delivered by CHWs with a person-centered contraceptive counseling component in a low-resource setting where contraception is underutilized. Our findings suggest the intervention’s potential to increase in modern contraceptive uptake in community settings. Perhaps more importantly, our results underscore the importance of longitudinal engagement and trust-building that position CHWs to drive longer-term change in their communities especially around sensitive topics. This study contributes to the implementation research literature on community-based interventions to improve postpartum contraception use and may inform other CHW programs in similar contexts.

## Data Availability

De-identified, ward-level data used for the primary analysis are publicly available at: https://osf.io/24smk/. Requests for participant level (human subjects) data can be sent to the corresponding author.

## References

1. Galati AJ. Onward to 2030: Sexual and Reproductive Health and Rights in the Context of the Sustainable Development Goals. 2015;18(4):8.

2. Starbird E, Norton M, Marcus R. Investing in Family Planning: Key to Achieving the Sustainable Development Goals. Global Health: Science and Practice. 2016;4(2):191–210.

3. Hardee K, Harris S, Rodriguez M, Kumar J, Bakamjian L, Newman K, et al. Achieving the goal of the London Summit on Family Planning by adhering to voluntary, rights-based family planning: what can we learn from past experiences with coercion? International perspectives on sexual and reproductive health. 2014;40(4):206–14.

4. United Nations Population Fund. Family Planning [Available from: https://www.unfpa.org/family-planning#readmore-expand.

5. Conde-Agudelo A, Rosas-Bermúdez A, Kafury-Goeta AC. Birth spacing and risk of adverse perinatal outcomes: a meta-analysis. JAMA. 2006;295(15):1809–23.

6. World Health Organization. Report of a WHO technical consultation on birth spacing: Geneva, Switzerland 13-15 June 2005. World Health Organization; 2007.

7. Moore Z, Pfitzer A, Gubin R, Charurat E, Elliott L, Croft T. Missed opportunities for family planning: an analysis of pregnancy risk and contraceptive method use among postpartum women in 21 low– and middle-income countries. Contraception. 2015;92(1):31–9.

8. Dev R, Kohler P, Feder M, Unger JA, Woods NF, Drake AL. A systematic review and meta-analysis of postpartum contraceptive use among women in low– and middle-income countries. Reproductive Health. 2019;16(1):154.

9. Gilmore K, Gebreyesus TA. What will it take to eliminate preventable maternal deaths? The Lancet. 2012;380(9837):87-8.

10. Ahmed S, Li Q, Liu L, Tsui AO. Maternal deaths averted by contraceptive use: an analysis of 172 countries. The Lancet. 2012;380(9837):111–25.

11. Government of Nepal Ministry of Health and Population. Nepal Demographic and Health Survey Key Indicators Report. 2022.

12. Puri MC, Huber-Krum S, Canning D, Guo M, Shah IH. Does family planning counseling reduce unmet need for modern contraception among postpartum women: Evidence from a stepped-wedge cluster randomized trial in Nepal. PLOS ONE. 2021;16(3):e0249106.

13. Mehata S, Paudel YR, Mehta R, Dariang M, Poudel P, Barnett S. Unmet need for family planning in Nepal during the first two years postpartum. Biomed Res Int. 2014;2014.

14. Pant PD, Pandey JP, Bietsch K. Unmet need for family planning and fertility in Nepal: levels, trends, and determinants. DHS Further Analysis Report. 2019(119).

15. Puri MC, Moroni M, Pearson E, Pradhan E, Shah IH. Investigating the quality of family planning counselling as part of routine antenatal care and its effect on intended postpartum contraceptive method choice among women in Nepal. BMC Women’s Health. 2020;20:29.

16. Ministry of Health NNEaI. Nepal Demographic and Health Survey 2016. Kathmandu, Nepal; 2017.

17. Maru DS-R, Andrews J, Schwarz D, Schwarz R, Acharya B, Ramaiya A, et al. Crossing the quality chasm in resource-limited settings. Global Health. 2012;8:41.

18. Mehata S, Paudel YR, Dariang M, Aryal KK, Lal BK, Khanal MN, et al. Trends and Inequalities in Use of Maternal Health Care Services in Nepal: Strategy in the Search for Improvements. Biomed Res Int. 2017;2017:5079234.

19. Khatri RB, Mishra SR, Khanal V. Female Community Health Volunteers in Community-Based Health Programs of Nepal: Future Perspective. Front Public Health. 2017;5:181.

20. Schwarz D, Sharma R, Bashyal C, Schwarz R, Baruwal A, Karelas G, et al. Strengthening Nepal’s Female Community Health Volunteer network: a qualitative study of experiences at two years. BMC Health Services Research. 2014;14(1):473.

21. Nepal P, Schwarz R, Citrin D, Thapa A, Acharya B, Acharya Y, et al. Costing Analysis of a Pilot Community Health Worker Program in Rural Nepal. Global Health: Science and Practice. 2020;8(2):239–55.

22. Organization WH. What do we know about community health workers? A systematic review of existing reviews. 2020.

23. Division FW, Population MoHa, Government of Nepal. NEPAL SAFE MOTHERHOOD AND NEWBORN HEALTH ROAD MAP 2030 2019.

24. World Health Organization. WHO guideline on health policy and system support to optimize community health worker programmes. 2018.

25. Maru S, Nirola I, Thapa A, Thapa P, Kunwar L, Wu W-J, et al. An integrated community health worker intervention in rural Nepal: a type 2 hybrid effectiveness-implementation study protocol. Implementation Science. 2018;13(1):53.

26. Tiwari A, Thapa A, Choudhury N, Khatri R, Sapkota S, Wu W-J, et al. A Type II hybrid effectiveness-implementation study of an integrated CHW intervention to address maternal healthcare in rural Nepal. PLOS Global Public Health. 2023;3(1):e0001512.

27. Gaffield ME, Egan S, Temmerman M. It’s about time: WHO and partners release programming strategies for postpartum family planning. Global Health: Science and Practice. 2014;2(1):4–9.

28. Cleland J, Shah IH, Daniele M. Interventions to Improve Postpartum Family Planning in Low– and Middle-Income Countries: Program Implications and Research Priorities. Studies in Family Planning. 2015;46(4):423–41.

29. Wu W-J, Tiwari A, Choudhury N, Basnett I, Bhatt R, Citrin D, et al. Community-based postpartum contraceptive counselling in rural Nepal: a mixed-methods evaluation. Sex Reprod Health Matters. 2020;28(2):1765646.

30. Curran GM, Bauer M, Mittman B, Pyne JM, Stetler C. Effectiveness-implementation Hybrid Designs. Med Care. 2012;50(3):217–26.

31. Glasgow RE, Vogt TM, Boles SM. Evaluating the public health impact of health promotion interventions: the RE-AIM framework. American journal of public health. 1999;89(9):1322–7.

32. Glasgow RE, Harden SM, Gaglio B, Rabin B, Smith ML, Porter GC, et al. RE-AIM planning and evaluation framework: adapting to new science and practice with a 20-year review. Front Public Health. 2019;7:64.

33. Vaidya NK, Wu J. HIV epidemic in Far-Western Nepal: effect of seasonal labor migration to India. BMC Public Health. 2011;11:310.

34. Citrin D, Thapa P, Nirola I, Pandey S, Kunwar LB, Tenpa J, et al. Developing and deploying a community healthcare worker-driven, digitally-enabled integrated care system for municipalities in rural Nepal. Healthc (Amst). 2018;6(3):197–204.

35. Dehlendorf C, Grumbach K, Schmittdiel JA, Steinauer J. Shared decision making in contraceptive counseling. Contraception. 2017;95(5):452–5.

36. Dehlendorf C, Krajewski C, Borrero S. Contraceptive counseling: best practices to ensure quality communication and enable effective contraceptive use. Clinical obstetrics and gynecology. 2014;57(4):659.

37. Holt K, Zavala I, Quintero X, Mendoza D, McCormick MC, Dehlendorf C, et al. Women’s preferences for contraceptive counseling in Mexico: results from a focus group study. Reproductive Health. 2018;15:1–11.

38. Agarwal S, Lasway C, L’Engle K, Homan R, Layer E, Ollis S, et al. Family Planning Counseling in Your Pocket: A Mobile Job Aid for Community Health Workers in Tanzania. Global Health: Science and Practice. 2016;4(2):300–10.

39. Population C. The Balanced Counseling Strategy Plus: A Toolkit for Family Planning Service Providers Working in High HIV/STI Prevalence Settings. Reproductive Health. 2015.

40. Liambila W, Warren C, Mullick S, Askew I, Homan R, Mohammed I, et al. Feasibility, acceptability, effect, and cost of integrating counseling and testing for HIV within family planning services in Kenya. Reproductive Health. 2008.

41. Sapkota S, Rajbhandary R, Lohani S. The Impact of Balanced Counseling on Contraceptive Method Choice and Determinants of Long Acting and Reversible Contraceptive Continuation in Nepal. Matern Child Health J. 2017;21(9):1713–23.

42. Government of Nepal Ministry of Health and Population Department of Health Services Nursing and Social Security Division. National Guideline on Community Health Program 2078. 2021.

43. Baqui AH, Ahmed S, Begum N, Khanam R, Mohan D, Harrison M, et al. Impact of integrating a postpartum family planning program into a community-based maternal and newborn health program on birth spacing and preterm birth in rural Bangladesh. J Glob Health. 2018;8(2):020406.

44. Ahmed S, Ahmed S, McKaig C, Begum N, Mungia J, Norton M, et al. The effect of integrating family planning with a maternal and newborn health program on postpartum contraceptive use and optimal birth spacing in rural Bangladesh. Studies in Family Planning. 2015;46(3):297–312.

45. Keogh SC, Urassa M, Kumogola Y, Kalongoji S, Kimaro D, Zaba B. Postpartum Contraception in Northern Tanzania: Patterns of Use, Relationship to Antenatal Intentions, and Impact of Antenatal Counseling. Studies in Family Planning. 2015;46(4):405–22.

46. Joshi AK, Tiwari DP, Poudyal A, Shrestha N, Acharya U, Dhungana GP. Utilization of family planning methods among postpartum mothers in Kailali district, Nepal. International journal of women’s health. 2020:487–94.

47. Staveteig S, Shrestha N, Gurung S, Kampa KT. Barriers to family planning use in Eastern Nepal: Results from a mixed methods study. 2018. Contract No.: 21.

48. Shattuck D, Wasti SP, Limbu N, Chipanta NS, Riley C. Men on the move and the wives left behind: the impact of migration on family planning in Nepal. Sex Reprod Health Matters. 2019;27(1):248–61.

49. Hendrickson ZM, Owczarzak J, Lohani S, Thapaliya Shrestha B, Underwood CR. The (re) productive work of labour migration: the reproductive lives of women with an absent spouse in the central hill region of Nepal. Culture, health & sexuality. 2019;21(6):684–700.

50. Mumah JN, Machiyama K, Mutua M, Kabiru CW, Cleland J. Contraceptive adoption, discontinuation, and switching among postpartum women in Nairobi’s urban slums. Studies in family planning. 2015;46(4):369–86.

51. Dehlendorf C, Reed R, Fox E, Seidman D, Hall C, Steinauer J. Ensuring our research reflects our values: The role of family planning research in advancing reproductive autonomy. Contraception. 2018;98(1):4–7.

52. Senderowicz L. “I was obligated to accept”: A qualitative exploration of contraceptive coercion. Soc Sci Med. 2019;239:112531.

53. Bullington BW, Sawadogo N, Tumlinson K, Langer A, Soura A, Zabre P, et al. Prevalence of non-preferred family planning methods among reproductive-aged women in Burkina Faso: results from a cross-sectional, population-based study. Sex Reprod Health Matters. 2023;31(1):2174244.

54. Bhatt N, Bhatt B, Neupane B, Karki A, Bhatta T, Thapa J, et al. Perceptions of family planning services and its key barriers among adolescents and young people in Eastern Nepal: A qualitative study. PLOS ONE. 2021;16(5):e0252184.

55. Sigdel A, Bista A, Sapkota H, Teijlingen Ev. Barriers in accessing family planning services in Nepal during the COVID-19 pandemic: A qualitative study. PLOS ONE. 2023;18(5):e0285248.

